# Associations of a multidimensional polygenic sleep health score and a sleep lifestyle index on health outcomes and their interaction in a clinical biobank

**DOI:** 10.1101/2024.02.06.24302416

**Authors:** Valentina Paz, Hannah Wilcox, Matthew Goodman, Heming Wang, Victoria Garfield, Richa Saxena, Hassan S. Dashti

**Affiliations:** Instituto de Psicología Clínica, Facultad de Psicología, Universidad de la República, Montevideo, Uruguay; MRC Unit for Lifelong Health & Ageing, Institute of Cardiovascular Science, University College London, London, United Kingdom; Center for Genomic Medicine, Massachusetts General Hospital and Harvard Medical School, Boston, Massachusetts, United States of America; Department of Anesthesia, Critical Care and Pain Medicine, Massachusetts General Hospital and Harvard Medical School, Boston, Massachusetts, United States of America; Brigham and Women’s Hospital, Harvard Medical School, Boston, Massachusetts, United States of America; Broad Institute, Cambridge, Massachusetts, United States of America; Division of Sleep Medicine, Harvard Medical School, Boston, Massachusetts, United States of America; Division of Nutrition, Harvard Medical School, Boston, Massachusetts, United States of America

**Keywords:** Multidimensional sleep, Polygenic scores, Lifestyle behaviors, Phenome-wide association study, Clinical disorders, Mental health

## Abstract

Sleep is a complex behavior regulated by genetic and environmental factors, and is known to influence health outcomes. However, the effect of multidimensional sleep encompassing several sleep dimensions on diseases has yet to be fully elucidated. Using the Mass General Brigham Biobank, we aimed to examine the association of multidimensional sleep with health outcomes and investigate whether sleep behaviors modulate genetic predisposition to unfavorable sleep on mental health outcomes. First, we generated a Polygenic Sleep Health Score using previously identified single nucleotide polymorphisms for sleep health and constructed a Sleep Lifestyle Index using data from self-reported sleep questions and electronic health records; second, we performed phenome-wide association analyses between these indexes and clinical phenotypes; and third, we analyzed the interaction between the indexes on prevalent mental health outcomes. Fifteen thousand eight hundred and eighty-four participants were included in the analysis (mean age 54.4; 58.6% female). The Polygenic Sleep Health Score was associated with the Sleep Lifestyle Index (β=0.050, 95%CI=0.032, 0.068) and with 114 disease outcomes spanning 12 disease groups, including obesity, sleep, and substance use disease outcomes (p<3.3×10^−5^). The Sleep Lifestyle Index was associated with 458 disease outcomes spanning 17 groups, including sleep, mood, and anxiety disease outcomes (p<5.1×10^−5^). No interactions were found between the indexes on prevalent mental health outcomes. These findings suggest that favorable sleep behaviors and genetic predisposition to healthy sleep may independently be protective of disease outcomes. This work provides novel insights into the role of multidimensional sleep on population health and highlights the need to develop prevention strategies focused on healthy sleep habits.

## Introduction

Sleep disturbance has numerous adverse health consequences, including psychiatric and cardiometabolic disorders [1]. The impact of sleep on health outcomes has predominantly been studied by considering individual aspects of sleep, such as duration or timing separately. However, a shift in recent years has emphasized sleep as a multidimensional construct and operationalized sleep health as a composite measure encompassing several different aspects. Combining individual sleep characteristics effectively generates an index reflecting overall ‘good’ or ‘bad’ sleep [2–4]. Several recent studies have used composite sleep metrics to investigate the relationship between sleep and health outcomes. These include studies reporting associations between ‘unhealthy’ sleep scores, derived from self-report and actigraphy, and more depressive symptoms [5] and higher risk of cardiovascular conditions [6], and between ‘healthy’ sleep scores and better mental well-being [7], cardiometabolic health [8], and lower psychological distress [9].

Sleep is a complex behavior regulated in part by environmental and genetic factors [10,11]. Recent genome-wide association studies (GWAS) have identified genetic variants robustly associated with composite sleep [12–14], enabling the computation of Polygenic Scores (PRS) (predictors of genetic susceptibility to traits or diseases) of multidimensional sleep [15]. Combining PRS with phenotypic risk factors could help elucidate the interplay between genetic and environmental influences on disease risk [16,17]. This inquiry is essential due to the large role of behavior and environmental constraints in sleep health in addition to sleep genetics [18]. Whether sleep behaviors can modulate genetic predispositions remains poorly understood.

Large clinical biobanks combine electronic health records (EHR) with genetic data and health surveys, providing resources to systematically interrogate genetic and lifestyle factors that influence multidimensional sleep and their relationship with hundreds of clinical phenotypes through phenome-wide scans [19–21]. In the present study, we leveraged the Mass General Brigham (MGB) Biobank to examine the association of multidimensional sleep (based on composite metrics of phenotypic and genetic data) with health outcomes and to investigate whether sleep behaviors can modulate sleep-related genetic predisposition. To achieve this, we (1) calculated a Polygenic Sleep Health Score for each participant based on GWAS of a composite sleep health score, (2) constructed an index of ‘healthy’ sleep based on data from self-reported questions regarding sleep patterns and EHR, and calculated a phenotypic Sleep Lifestyle Index for each participant, (3) conducted a hypothesis-free phenome-wide association analysis (PheWAS) to identify health outcomes associated with the genetic and phenotypic scores, and (4) analyzed the interaction between the Polygenic Sleep Health Score and the Sleep Lifestyle Index on the ‘top hits’ for mental health outcomes from the PheWAS.

## Methods

### Sample

Study participants were patients from the Mass General Brigham (MGB) Biobank. The MGB Biobank is a healthcare enterprise clinical cohort from the MGB healthcare network in Massachusetts. The MGB Biobank links EHR with genetic and lifestyle data. Since 2009, patients have been recruited online through the patient portal or in person across multiple MGB community-based primary care facilities and specialty tertiary care centers [20,22,23]. The recruitment strategy has been described previously [20]. Written informed consent was obtained from all patients upon enrollment (Spanish translation was available to promote patient inclusivity). The present study protocol was approved by the MGB Institutional Review Board (#2018P002276). At the time of the present analysis (03/2023), 140,915 patients had enrolled in the biobank.

### Polygenic Sleep Health Score

Among the enrolled patients, 64,639 patients (45.9% of the total) had provided blood samples available for genotyping. DNA from samples was genotyped using the Infinium Global Screening Array-24 version 2.0 (Illumina). Imputation was performed using the Michigan Imputation server with the Trans-Omics for Precision Medicine (TOPMed) (version r2) reference panel [24], and haplotype phasing was performed using Eagle version 2.3. As previously described, the genetic data were quality controlled, excluding low-quality genetic markers and samples [25]. Pairs of related individuals (kinship > 0.0625) were identified, and one sample from each related pair was excluded. Using TRACE and the Human Genome Diversity Project, principal components of ancestry were computed to correct for the population substructure [26,27]. Participants of non-European ancestry (24.9% of the cohort) were excluded from the analysis to limit genetic heterogeneity in the present study.

Effect estimates for the Polygenic Sleep Health Score were derived from the most recent large-scale GWAS of composite sleep in UK Biobank participants of European ancestry (n=413,904) [14]. In this GWAS, a clinical additive sleep health score was derived from five favorable binary attributes of underlying ordinal sleep traits from self-report (7-8 hours sleep duration, early chronotype, few insomnia symptoms, no snoring, and no excessive daytime sleepiness) [28]. For each participant in the present MGB Biobank study, a composite Polygenic Sleep Health Score was generated using Polygenic Risk Score–Continuous Shrinkage (PRS-CS) [29]. This method is based on Bayesian regression and places a continuous shrinkage prior on single-nucleotide polymorphism effect sizes. The UKBiobank European ancestry linkage disequilibrium (LD) panel was used for LD pruning. A total of 239,446 single-nucleotide polymorphisms were included in the score following clumping. The score was standardized with a mean of 0 and a standard deviation (SD) of 1.

### Sleep Lifestyle Index

All participants enrolled in the MGB Biobank were invited to complete an optional Health Information Questionnaire (HIQ) composed of lifestyle and family history questions. The following four questions related to sleep habits were asked with responses in half-hour increments: “In considering your longest sleep period, what time do you usually go to bed on weekdays or work or school days?” and “In considering your longest sleep period, what time do you usually wake up on weekdays or work or school days?” (both also asked for “weekends or days off”). At the time of analysis, 65,248 (46.30%) participants responded to the optional questionnaire. Self-reported bedtimes between 08:00 am and 02:00 pm (weekday *n*=329; weekend *n*=613) and self-reported wake times between 06:00 pm and 12:00 am (weekday *n*=141; weekend *n*=287), likely resulting from am/pm misreporting, were set to missing. Improbable time in bed <3 or >18 hours (weekday *n*=73; weekend *n*=74) were set to missing, consistent with previous analyses [30].

Using responses from these questions, the following was calculated: (1) time in bed as the weighted average weekly time in bed with 5/7 weighting for weekdays and 2/7 for weekends; (2) time in bed irregularity as the absolute value of the difference between weekday and weekend time in bed; (3) sleep midpoint as the midpoint of bed and wake times on weekends and; (4) social jetlag as the absolute difference in weekend and weekday sleep midpoint. From EHR data, sleep medications and sleep disorders, including insomnia and sleep apnea, based on the International Classification of Diseases (ICD)-9/-10 billing codes were determined. Participants with at least two codes for the same diagnosis on two separate dates within five years of completing the HIQ (to support cross-sectional analyses) were considered to have a sleep disorder, and those with no relevant code within this five-year time window were considered free of sleep disorders (those with only one code were set to missing).

A Sleep Lifestyle Index was constructed based on comparable attributes considered in the UK Biobank GWAS for a clinical additive sleep health score and other sleep indices based on available data [6,31]. The index aggregated exposure to each of the following sleep behaviors: (1) adequate time in bed (*≥*7 h and *≤*9 h per night) [32,33]; (2) regular time in bed (difference <60 min between weekday and weekend time in bed) [6]; (3) healthy sleep midpoint (between 2:00 am and 4:00 am) [31]; (4) absence or mild/moderate social jetlag (<2 h) [34]; (5) not taking any medication known to affect sleep, such as those used to treat insomnia, anxiety or circadian disorders (the list of medication is shown in Supplementary Material Table 1); (6) no recent diagnosis of any insomnia-related disorders; (7) no recent diagnosis of any sleep-related breathing disorders; and (8) no recent diagnosis of any other sleep disorder (the list of disorders is shown in Supplementary Material Table 2). Participants were assigned one point for each healthy sleep behavior [2]. The Sleep Lifestyle Index scores ranged from 0 to 8, with higher scores reflecting more favorable (less problematic) sleep behaviors. Cross-trait correlations are presented in Supplementary Material Table 3. The associations between the index and each sleep behavior are presented in Supplementary Material Table 4.

### Clinical outcomes

The clinical phenotypes were determined from the ICD-9/-10 billing codes in the EHR [20]. Codes were mapped to 1,846 phenome-wide association study (PheWAS) codes (i.e., clinical phenotypes “phecodes”) based on clinical similarity. Same-day duplicated diagnoses and non-ICD-9/-10 codes were removed. Participants with at least two codes for a disorder within five years of completing the HIQ were considered cases, and those with no relevant codes were considered controls [21,35].

### Statistical analysis

We tested associations between the Polygenic Sleep Health Score and the Sleep Lifestyle Index, and between the Polygenic Sleep Health Score and each sleep attribute included in the Sleep Lifestyle Index using linear or logistic regression models adjusted for age, sex, genotyping array, batch, and principal components of ancestry (primary model) and further adjusted for employment, education, exercise, smoking, alcohol intake, body mass index, and Charlson Comorbidity Index [36] (fully adjusted model). The description of the covariates is shown in Supplementary Material Table 5.

We conducted a PheWAS for the Polygenic Sleep Health Score with 1,354 disorders (with at least 100 cases in the analytical sample) in 47,082 unrelated adult participants of European ancestry with high-quality genetic data using the PheWAS R package [19]. We also conducted a PheWAS for the Sleep Lifestyle Index with 898 disorders in 15,884 patients (33.7% of genotyped European patients) diagnosed within five years of completing the HIQ.

We tested associations between the Polygenic Sleep Health Score and each disorder using logistic regressions adjusted for age, sex, genotyping array, batch, and principal components of ancestry. We tested associations between the Sleep Lifestyle Index and each disorder using logistic regressions adjusted for age, sex, employment, education, exercise, smoking, alcohol intake, body mass index, and the Charlson Comorbidity Index. Finally, we conducted interaction tests between the Polygenic Sleep Health Score and the Sleep Lifestyle Index for the top five mental health outcomes associated with both indexes in the PheWAS by further adding an interaction term between the indexes.

Significance was determined at Bonferroni P value cut-offs accounting for the total number of tests. Descriptive statistics are presented as mean ± standard deviation. All analyses were performed using R statistical computing (version 2022.12.0; The R Foundation for Statistical Computing, Vienna, Austria).

## Results

A total of 47,082 adult patients of European ancestry were included in the genetic analysis (mean age = 60.4 ± 17.0; 53.8% female), and 15,884 patients were included in the lifestyle analysis (mean age = 54.4 ± 16.3; 58.6% female) (Figure 1).

**Figure 1.**
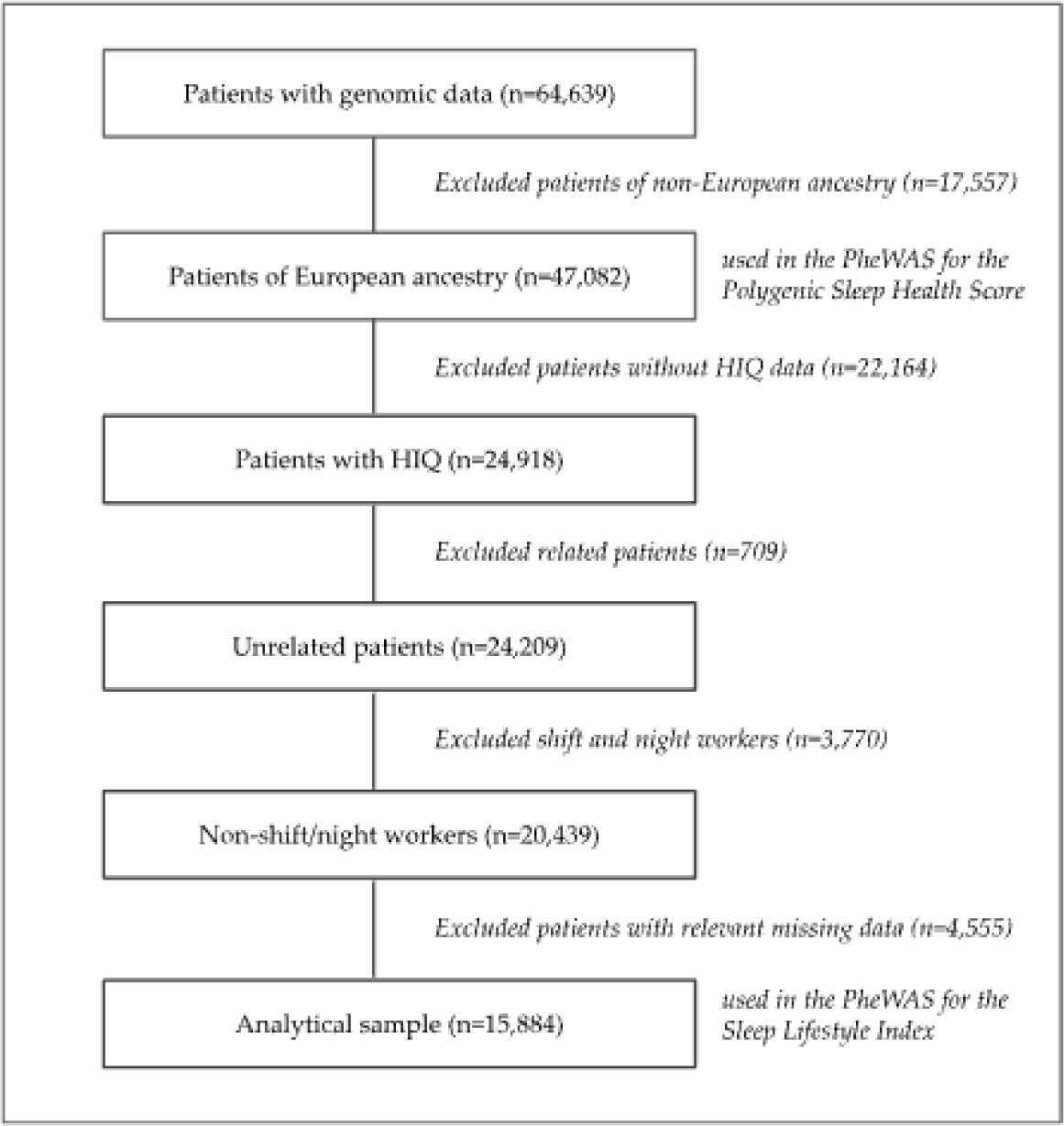
Flowchart of patients included in the analyses and exclusion criteria. HIQ: Health Information Questionnaire. PheWAS: Phenome-wide association analysis.

The cohort averages for time in bed was 8.15 ± 1.13 h, time in bed regularity was 0.78 ± 0.94 h (range: 0-12 h), sleep midpoint was 03:23 ± 01:41, and social jetlag was 0.86 ± 0.82 h (range: 0-10.5 h). The prevalence of insomnia-related disorders, sleep-related breathing disorders, and any other sleep disorders based on recent diagnoses were 1.9%, 3.5%, and 2.9%, respectively. All participants presented at least one of the healthy sleep lifestyle traits, while 4421 participants (27.83%) presented with all 8 traits. Most participants presented with at least half of the healthy sleep behaviors ascertained (adequate time in bed, time in bed regularity, healthy sleep midpoint, and mild social jetlag) (Figure 2).

**Figure 2.**
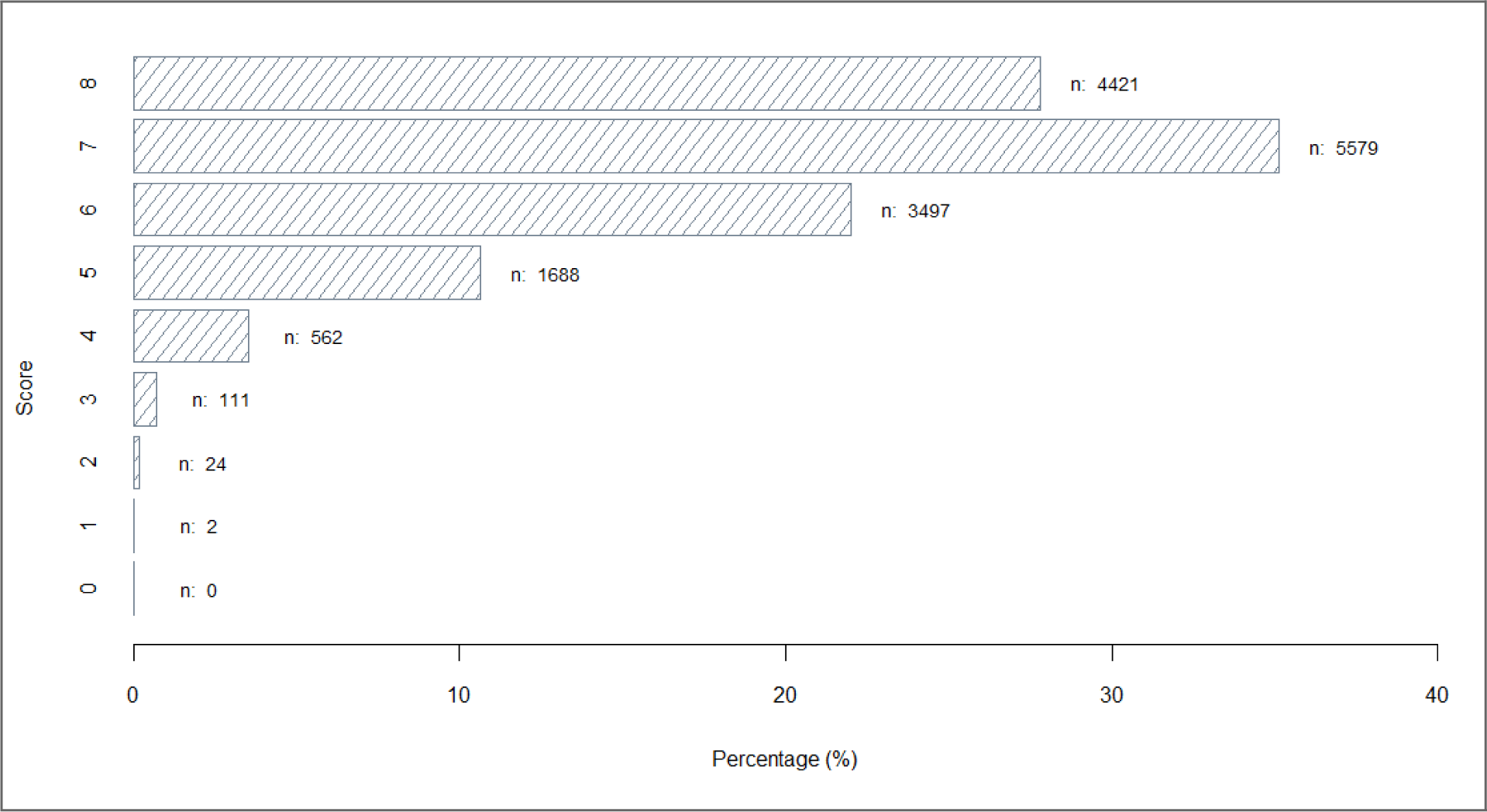
Histogram of Sleep Lifestyle Index scores based on self-reported sleep traits and data from electronic health records. The scores ranged from 0 to 8, with higher scores reflecting more favorable (less problematic) sleep behaviors.

The Polygenic Sleep Health Score was associated with the composite Sleep Lifestyle Index (primary model: β=0.077, 95%CI=0.058, 0.095; fully adjusted model: β=0.050, 95% CI=0.032, 0.068). Each SD increment in the Polygenic Sleep Health Score was associated with several components of the index, including less sleep irregularity (primary model: β=-0.026, 95% CI=-0.041, −0.011; fully adjusted model: β=-0.018, 95% CI=-0.033, −0.003), earlier sleep midpoint (primary model: β=-0.066, 95% CI=-0.093, −0.039; fully adjusted model: β=-0.057, 95% CI=-0.084, −0.030), less social jetlag (primary model: β=-0.022, 95% CI=-0.034, −0.010; fully adjusted model: β=-0.018, 95% CI=-0.030, −0.007) and, in the primary model only, higher odds of not having sleep-related breathing disorders (primary model: OR=1.165, 95% CI=1.065, 1.275; fully adjusted model: OR=1.075, 95% CI=0.980, 1.178) and higher odds of not having any other sleep disorder (primary model: OR=1.165, 95% CI=1.057, 1.285; fully adjusted model: OR=1.082, 95% CI=0.979, 1.196). The Polygenic Sleep Health Score was not associated with adequate time in bed, insomnia-related disorders, and medication (P value>0.05).

### PheWAS results

PheWAS results for the Polygenic Sleep Health Score are presented in Figure 3 and in Supplementary Material Table 6. Significant findings were evident for 114 disease outcomes spanning 12 disease groups (p<3.3×10^−5^). The Polygenic Sleep Health Score was negatively associated with mental health outcomes, accounting for 14.0% of all findings. The 10 strongest associations for mental health outcomes were Tobacco use disorder (OR=0.881, 95% CI=0.861, 0.902), Substance addiction and disorders (OR=0.858, 95% CI=0.828, 0.889), Major depressive disorder (OR=0.905, 95% CI=0.882, 0.929), Depression (OR=0.906, 95%CI=0.883, 0.930), Anxiety disorder (OR=0.916, 95% CI=0.893, 0.939), Alcoholism (OR=0.869, 95% CI=0.833, 0.908), Posttraumatic stress disorder (OR=0.848, 95% CI=0.806, 0.892), Alcohol-related disorders (OR=0.888, 95% CI=0.850, 0.928), Mood disorders (OR=0.904, 95% CI=0.871, 0.939) and, Adjustment reaction (OR=0.924, 95% CI=0.897, 0.952) (Figure 5) (results were similar when adjusting for body mass index (BMI), see Supplementary Material Table 7).

**Figure 3.**
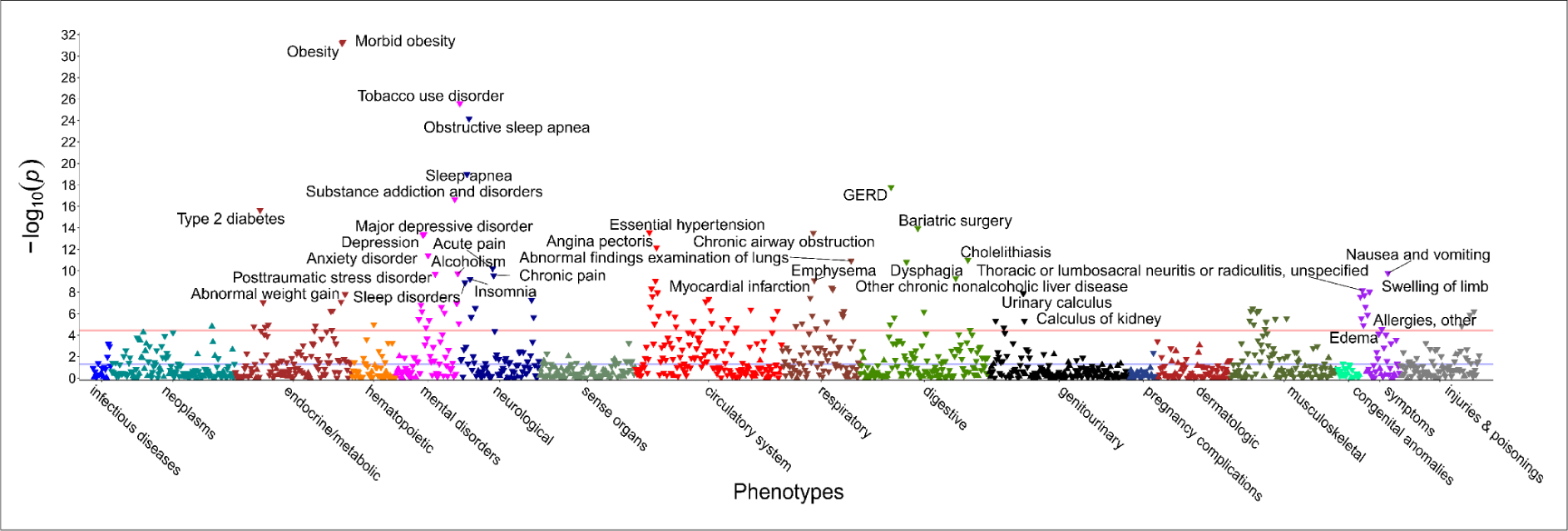
Manhattan plot of phenome-wide associations between the Polygenic Sleep Health Score and disease outcomes (grouped by broad disease groups), in a model adjusting for age, sex, genotyping array, batch, and principal components of ancestry. The -log10P value of the association is shown on the y-axis. The horizontal lines represent Bonferroni corrected P value cut-offs (the red line represents P value = 5 × 10^−5^; the blue line represents P value = 8 × 10^−4^). Upward triangles indicate positive associations, and downward triangles indicate negative associations.

PheWAS results for the Sleep Lifestyle Index are presented in Figure 4 and in Supplementary Material Table 8. Significant findings were evident for 458 disease outcomes spanning 17 disease groups (p<5.1×10^−5^). Similarly, the Sleep Lifestyle Index was negatively associated with mental health outcomes for 9.0% of significant findings. The 10 strongest associations for mental health outcomes were Depression (OR=0.668, 95% CI=0.640, 0.696), Major depressive disorder (OR=0.683, 95% CI=0.655, 0.711), Anxiety disorder (OR=0.693, 95% CI=0.666, 0.721), Generalized anxiety disorder (OR=0.662, 95% CI=0.626, 0.700), Mood disorders (OR=0.630, 95% CI=0.588, 0.675), Adjustment reaction (OR=0.719, 95% CI=0.683, 0.756), Altered mental status (OR=0.654, 95% CI=0.612, 0.698), Dysthymic disorder (OR=0.550, 95% CI=0.500, 0.605), Bipolar (OR=0.641, 95% CI=0.593, 0.692) and, Memory loss (OR=0.676, 95% CI=0.632, 0.724) (Figure 5).

**Figure 4.**
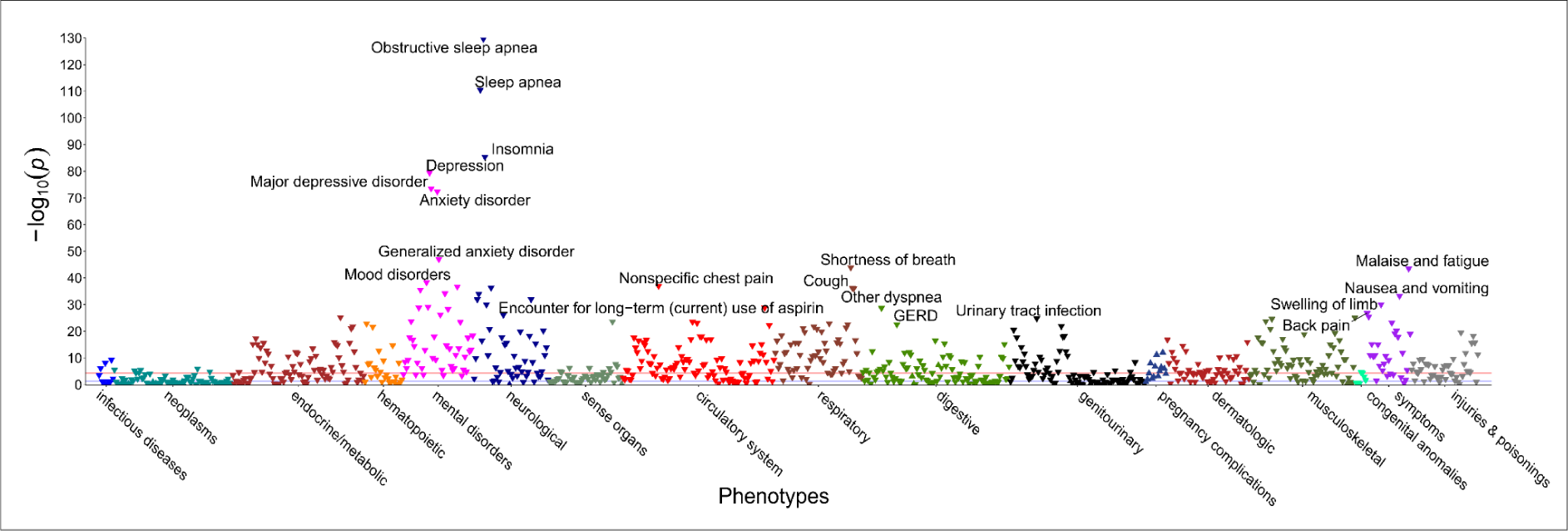
Manhattan plot of phenome-wide associations between the Sleep Lifestyle Index and disease outcomes (grouped by broad disease groups), in a model adjusting for age, sex, employment, education, exercise, smoking, alcohol intake, body mass index, and the Charlson Comorbidity Index. The -log10P value of the association is shown on the y-axis. The horizontal lines represent Bonferroni corrected P value cut-offs (the red line represents P value = 5 × 10^−5^; the blue line represents P value = 8 × 10^−4^). Upward triangles indicate positive associations, and downward triangles indicate negative associations.

**Figure 5.**
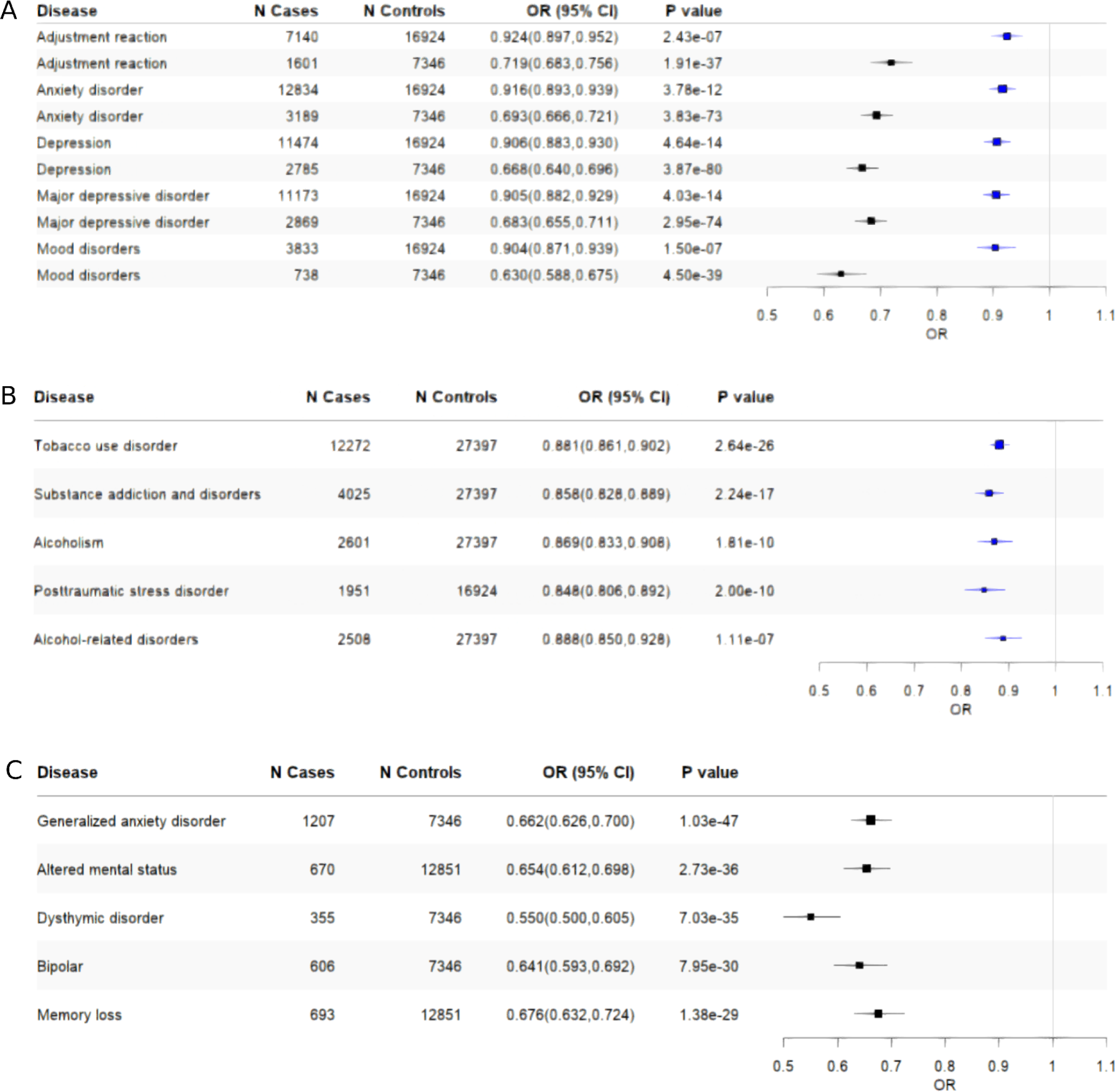
A) Phenome-wide associations between the Polygenic Sleep Health Score (blue) and the Sleep Lifestyle Index (black) and the five mental health outcomes strongly associated with both indexes. B) Phenome-wide associations between the Polygenic Sleep Health Score and the other five mental health outcomes strongly associated with the index. C) Phenome-wide associations between the Sleep Lifestyle Index and the other five mental health outcomes strongly associated with the index. Notes: OR, Odd ratio; 95% CI, 95% confidence interval.

Taking into account that five mental health outcomes were strongly associated with both indexes (Figure 5 A, see description in Supplementary Material 9) and previous work linking multidimensional sleep with mental health [5,7,9], we investigated whether these indexes interact to explain them. No significant interactions were observed between the indexes on these mental health outcomes (all P > 0.05) (Table 1).

**Table 1.** Interactions between the Polygenic Sleep Health Score and the Sleep Lifestyle Index on disease outcomes. Disease outcomes were limited to the five mental health outcomes strongly associated with both indexes. Notes: OR, odd ratio; 95% CI, 95% confidence interval.

| Disease | Primary model |  | Fully adjusted model |  |
| --- | --- | --- | --- | --- |
|  | OR (95% CI) | P-value | OR (95% CI) | P-value |
| Adjustment reaction | 0.984 (0.935, 1.036) | 0.523 | 0.983 (0.934, 1.035) | 0.501 |
| Anxiety disorder | 0.992 (0.954, 1.032) | 0.704 | 0.988 (0.948, 1.030) | 0.558 |
| Depression | 1.004 (0.963, 1.046) | 0.863 | 1.002 (0.960, 1.046) | 0.918 |
| Major depressive disorder | 0.996 (0.956, 1.038) | 0.843 | 0.994 (0.954, 1.036) | 0.795 |
| Mood disorders | 0.986 (0.921, 1.056) | 0.696 | 0.975 (0.909, 1.047) | 0.491 |

## Discussion

In the present study, we examined the link between behavioral and genetic multidimensional sleep indexes with health outcomes and investigated whether sleep behaviors can modulate genetic predisposition. To achieve this, we calculated, for the first time, a polygenic score for composite sleep and constructed a lifestyle index based on sleep behaviors. Both indexes were associated with each other and with health outcomes, including mental, neurological, and endocrine/metabolic disorders. However, significant evidence of the specified parametric interactions between the indexes was not observed for prevalent mental health conditions suggesting that improving sleep habits can mitigate the risk of disease irrespective of genetic predisposition.

We found that each standard deviation increment in the genetic index was associated with a 0.050-0.077 unit increase (0.625–0.963%) in the lifestyle index. Previous studies have reported associations between polygenic scores for self-reported sleep duration, insomnia, and chronotype with their corresponding traits [37–40]. Thus, our finding contributes to validating sleep polygenic scores as proxies for sleep behaviors, including multidimensional sleep. Moreover, we found that the genetic index was consistently negatively associated with disease outcomes, suggesting that the genetic predisposition to healthy sleep may be protective against multiple disorders; however, a causal role for sleep in these disorders cannot be inferred from the present analyses. Interestingly, the lifestyle index was also strongly associated with several diseases, which is consistent with the role of healthy sleep behaviors in improving overall health and the benefit of routine assessment of sleep disturbances in clinical services. In this line, two recent systematic reviews have reported that unhealthy sleep patterns, such as short and long sleep durations, high sleep variability, and late sleep timing, are associated with adverse health outcomes [41,42].

Mental disorders were strongly associated with both indexes, particularly mood, anxiety, and reaction to severe stress and adjustment disorders. Specifically, the genetic index was associated with 8-10% lower odds of having these disorders, while the lifestyle index was associated with 28-37% lower odds. Noteworthily, these disorders are the most frequently diagnosed psychiatric conditions [43,44]. The link between sleep difficulties and psychiatric illness is well recognized; however, the effect of sleep on mental health is not fully understood. Current evidence suggests a causal role of sleep behaviors in developing these disorders [45]. For example, Mendelian randomization (MR) studies have found a decreased risk of depression in individuals with morning chronotype [46,47], an increased risk of anxiety in those with insomnia [48], and bidirectional associations between insomnia and depression [49,50]. Further, a recent study has reported substantial polygenic overlap between sleep-related traits and some mental conditions, including depression [51]. Moreover, the GWAS used to generate the genetic index reported genetic correlations between the composite sleep health score and several mental health conditions, including mental distress, anxiety, and depression [28]. In addition, a meta-analysis of randomized controlled trials found that improving sleep quality reduces depression and anxiety symptoms [52]. Nonetheless, some MR studies did not find causal effects between chronotype or insomnia on depression [53,54]. This knowledge emphasizes the importance of thoroughly investigating the connection between sleep and mental health, offering new possibilities for therapeutic interventions.

Metabolic diseases were also associated with the indexes. Particularly, obesity was the phenotype most strongly associated with the genetic index. The link between sleep and obesity has been extensively reported [55,56]. Previous meta-analyses have revealed that sleep durations outside the normal range increase the incidence of obesity [57–59]. In agreement, prior research has reported U-shaped positive genetic correlations between short and long sleep durations and BMI, waist circumference, and waist-to-hip ratio [30]. However, it remains elusive whether the association between sleep duration and BMI is causal [60]. Sleep breathing disorders are also highly associated with obesity [61], and robust causal effects of insomnia on higher BMI have been reported in MR studies [62,63]. Regarding genetic risk, a previous study on the MGB Biobank indicated that a polygenic index for sleep duration was associated with obesity [37].

Finally, we did not find statistical interactions of genetic and lifestyle sleep factors on the five most prevalent mental health outcomes associated with both indexes. Recent work has shown that short/long sleep duration modifies genetic risk for adverse lipid profiles [64] and blood pressure [65]. Similarly, some gene-sleep interaction studies suggest favorable sleep behaviors may attenuate genetic predisposition to obesity [55]. However, for other phenotypes, such as type 2 diabetes, little compelling data supports gene-lifestyle interactions [66]. Thus, one possibility for our null findings is that, as for diabetes, lifestyle factors independently predict the mental health outcomes studied. It is also possible that we have inaccurate or incomplete patient diagnoses, which would contribute to the misclassification of cases. Nevertheless, the absence of evidence for the tested statistical interactions may suggest that improving sleep behaviors may ameliorate disease risk regardless of genetic background.

Our study has several strengths. First, unique to the MGB Biobank is the linking of large and diverse medical data enriched for disease with genetic and sleep information generally unavailable in other clinical biobanks. Second, using composite sleep metrics has several advantages, including the recognition that sleep dimensions do not occur in isolation and the possibility of analyzing gradients of healthy sleep beyond the absence/presence of sleep disturbances [67]. Third, composite metrics could enhance predictive power [4]. Finally, a score derived from genetic markers should act independently of confounders that could influence the associations between sleep and health outcomes.

Limitations of the study should also be acknowledged. The study was restricted to participants of European ancestry, as the discovery GWAS was conducted in Europeans [14]; future work in diverse racial and ethnic groups is necessary for the generalization of findings and to promote health equity. Moreover, our results cannot be generalized to other age groups and non-clinical cohorts. We focused on the MGB biobank; thus, diagnosis data is likely incomplete as we did not consider information from other medical facilities. Validating these indexes with diseases from additional electronic medical records is necessary. The exclusion of rare diseases (i.e., <100 cases) in the PheWAS analyses due to small case numbers warrants future exploration of multidimensional sleep effects on these disorders. Furthermore, our analyses were based on self-reported data. The modest HIQ response rate introduces potential selection bias, while the single survey administration limits insights into behavior stability over time. Lastly, given the cross-sectional nature of our study, as well as potential genetic pleiotropy, the reported associations do not necessarily imply causal relationships.

## Conclusions

The present study explored how genetic susceptibility to healthy sleep and beneficial sleep habits were associated with various disorders within a clinical cohort. While genetic and phenotypic sleep factors were linked to several diseases, no interactions were evident between these factors on mental health outcomes. Overall, our findings emphasize the relevance of sleep for a healthy life, demonstrate the pleiotropic nature of sleep genetics, and underscore the importance of leveraging clinical biobanks in advancing precision medicine research. Further research is needed on the association between multidimensional sleep and health outcomes in diverse clinical settings.

## Data Availability

Data are available from the Mass General Brigham Human Research Office/Institutional Review Board at Mass General Brigham (contact located at https://www.partners.org/Medical-Research/Support-Offices/Human-Research-Committee-IRB/Default.aspx) for researchers who meet the criteria for access to confidential data.

## Acknowledgments

We thank the participants and administrators of the MGB Biobank for their contribution to this work.

## References

1. Medic G, Wille M, Hemels M. Short- and long-term health consequences of sleep disruption. Nat Sci Sleep. 2017;Volume 9:151–161. doi:10.2147/NSS.S134864

2. Buysse DJ. Sleep Health: Can We Define It? Does It Matter? Sleep. 2014;37(1):9–17. doi:10.5665/sleep.3298

3. Matricciani L, Bin YS, Lallukka T, et al. Rethinking the sleep-health link. Sleep Health. 2018;4(4):339–348. doi:10.1016/j.sleh.2018.05.004

4. Wallace ML, Hall MH, Buysse DJ. Measuring sleep health. In: Foundations of Sleep Health. Elsevier; 2022:37–71. doi:10.1016/B978-0-12-815501-1.00015-6

5. Furihata R, Hall MH, Stone KL, et al. An Aggregate Measure of Sleep Health Is Associated With Prevalent and Incident Clinically Significant Depression Symptoms Among Community-Dwelling Older Women. Sleep. 2017;40(3). doi:10.1093/sleep/zsw075

6. Lee S, Smith CE, Wallace ML, et al. Cardiovascular risks and sociodemographic correlates of multidimensional sleep phenotypes in two samples of US adults. SLEEP Adv. 2022;3(1):zpac005. doi:10.1093/sleepadvances/zpac005

7. Appleton SL, Melaku YA, Reynolds AC, Gill TK, De Batlle J, Adams RJ. Multidimensional sleep health is associated with mental well-being in Australian adults. J Sleep Res. 2022;31(2). doi:10.1111/jsr.13477

8. Makarem N, Alcantara C, Musick S, et al. Multidimensional Sleep Health Is Associated with Cardiovascular Disease Prevalence and Cardiometabolic Health in US Adults. Int J Environ Res Public Health. 2022;19(17):10749. doi:10.3390/ijerph191710749

9. DeSantis AS, Dubowitz T, Ghosh-Dastidar B, et al. A preliminary study of a composite sleep health score: associations with psychological distress, body mass index, and physical functioning in a low-income African American community. Sleep Health. 2019;5(5):514–520. doi:10.1016/j.sleh.2019.05.001

10. Hale L, Emanuele E, James S. Recent Updates in the Social and Environmental Determinants of Sleep Health. Curr Sleep Med Rep. 2015;1(4):212–217. doi:10.1007/s40675-015-0023-y

11. Tafti M. Genetic aspects of normal and disturbed sleep. Sleep Med. 2009;10:S17–S21. doi:10.1016/j.sleep.2009.07.002

12. Lane JM, Liang J, Vlasac I, et al. Genome-wide association analyses of sleep disturbance traits identify new loci and highlight shared genetics with neuropsychiatric and metabolic traits. Nat Genet. 2017;49(2):274–281. doi:10.1038/ng.3749

13. Yao Y, Jia Y, Wen Y, et al. Genome-Wide Association Study and Genetic Correlation Scan Provide Insights into Its Genetic Architecture of Sleep Health Score in the UK Biobank Cohort. Nat Sci Sleep. 2022;Volume 14:1–12. doi:10.2147/NSS.S326818

14. Goodman M, Faquih T, Paz V, et al. Genome-Wide Association Analysis of Composite Sleep Health Scores in 413,904 Individuals. medRxiv; 2024. doi:10.1101/2024.02.02.24302211

15. Wray NR, Goddard ME, Visscher PM. Prediction of individual genetic risk to disease from genome-wide association studies. Genome Res. 2007;17(10):1520–1528. doi:10.1101/gr.6665407

16. Choi SW, Mak TSH, O’Reilly PF. Tutorial: a guide to performing polygenic risk score analyses. Nat Protoc. 2020;15(9):2759–2772. doi:10.1038/s41596-020-0353-1

17. Murray GK, Lin T, Austin J, McGrath JJ, Hickie IB, Wray NR. Could Polygenic Risk Scores Be Useful in Psychiatry?: A Review. JAMA Psychiatry. 2021;78(2):210. doi:10.1001/jamapsychiatry.2020.3042

18. Lopez R, Barateau L, Dauvilliers Y. [Normal organization of sleep and its changes during life]. Rev Prat. 2019;69(5):537–545.

19. Denny JC, Bastarache L, Roden DM. Phenome-Wide Association Studies as a Tool to Advance Precision Medicine. Annu Rev Genomics Hum Genet. 2016;17(1):353–373. doi:10.1146/annurev-genom-090314-024956

20. Karlson E, Boutin N, Hoffnagle A, Allen N. Building the Partners HealthCare Biobank at Partners Personalized Medicine: Informed Consent, Return of Research Results, Recruitment Lessons and Operational Considerations. J Pers Med. 2016;6(1):2. doi:10.3390/jpm6010002

21. Wei WQ, Bastarache LA, Carroll RJ, et al. Evaluating phecodes, clinical classification software, and ICD-9-CM codes for phenome-wide association studies in the electronic health record. Rzhetsky A, ed. PLOS ONE. 2017;12(7):e0175508. doi:10.1371/journal.pone.0175508

22. Boutin N, Mathieu K, Hoffnagle A, et al. Implementation of Electronic Consent at a Biobank: An Opportunity for Precision Medicine Research. J Pers Med. 2016;6(2):17. doi:10.3390/jpm6020017

23. Boutin N, Holzbach A, Mahanta L, et al. The Information Technology Infrastructure for the Translational Genomics Core and the Partners Biobank at Partners Personalized Medicine. J Pers Med. 2016;6(1):6. doi:10.3390/jpm6010006

24. Taliun D, Harris DN, Kessler MD, et al. Sequencing of 53,831 diverse genomes from the NHLBI TOPMed Program. Nature. 2021;590(7845):290–299. doi:10.1038/s41586-021-03205-y

25. Dashti HS, Hivert MF, Levy DE, McCurley JL, Saxena R, Thorndike AN. Polygenic risk score for obesity and the quality, quantity, and timing of workplace food purchases: A secondary analysis from the ChooseWell 365 randomized trial. Basu S, ed. PLOS Med. 2020;17(7):e1003219. doi:10.1371/journal.pmed.1003219

26. Cann HM, de Toma C, Cazes L, et al. A Human Genome Diversity Cell Line Panel. Science. 2002;296(5566):261–262. doi:10.1126/science.296.5566.261b

27. Wang C, Zhan X, Liang L, Abecasis GR, Lin X. Improved Ancestry Estimation for both Genotyping and Sequencing Data using Projection Procrustes Analysis and Genotype Imputation. Am J Hum Genet. 2015;96(6):926–937. doi:10.1016/j.ajhg.2015.04.018

28. Goodman M, Nagarajan P, Lane J, et al. 0033 Genome-wide association analysis of composite sleep scores in 413,904 individuals. SLEEP. 2023;46(Supplement_1):A15–A15. doi:10.1093/sleep/zsad077.0033

29. Ge T, Chen CY, Ni Y, Feng YCA, Smoller JW. Polygenic prediction via Bayesian regression and continuous shrinkage priors. Nat Commun. 2019;10(1):1776. doi:10.1038/s41467-019-09718-5

30. Dashti, Jones SE, Wood AR, et al. Genome-wide association study identifies genetic loci for self-reported habitual sleep duration supported by accelerometer-derived estimates. Nat Commun. 2019;10(1):1100. doi:10.1038/s41467-019-08917-4

31. Hawkins MS, Pokutnaya DY, Bodnar LM, et al. The Association between Multidimensional Sleep Health and Gestational Weight Gain: nuMoM2b Sleep Duration and Continuity Study. Obstetrics and Gynecology; 2023. doi:10.1101/2023.02.21.23285931

32. Hirshkowitz M, Whiton K, Albert SM, et al. National Sleep Foundation’s updated sleep duration recommendations: final report. Sleep Health. 2015;1(4):233–243. doi:10.1016/j.sleh.2015.10.004

33. Dashti HS, Cade BE, Stutaite G, Saxena R, Redline S, Karlson EW. Sleep health, diseases, and pain syndromes: findings from an electronic health record biobank. Sleep. 2021;44(3):zsaa189. doi:10.1093/sleep/zsaa189

34. Caliandro R, Streng AA, van Kerkhof LWM, van der Horst GTJ, Chaves I. Social Jetlag and Related Risks for Human Health: A Timely Review. Nutrients. 2021;13(12):4543. doi:10.3390/nu13124543

35. Wu P, Gifford A, Meng X, et al. Mapping ICD-10 and ICD-10-CM Codes to Phecodes: Workflow Development and Initial Evaluation. JMIR Med Inform. 2019;7(4):e14325. doi:10.2196/14325

36. Charlson ME, Pompei P, Ales KL, MacKenzie CR. A new method of classifying prognostic comorbidity in longitudinal studies: Development and validation. J Chronic Dis. 1987;40(5):373–383. doi:10.1016/0021-9681(87)90171-8

37. Dashti, Redline S, Saxena R. Polygenic risk score identifies associations between sleep duration and diseases determined from an electronic medical record biobank. Sleep. 2019;42(3). doi:10.1093/sleep/zsy247

38. Perkiö A, Merikanto I, Kantojärvi K, et al. Portability of Polygenic Risk Scores for Sleep Duration, Insomnia and Chronotype in 33,493 Individuals. Clocks Sleep. 2022;5(1):10–20. doi:10.3390/clockssleep5010002

39. Tsapanou A, Mourtzi N, Charisis S, et al. Sleep Polygenic Risk Score Is Associated with Cognitive Changes over Time. Genes. 2021;13(1):63. doi:10.3390/genes13010063

40. Tsapanou A, Gao Y, Stern Y, Barral S. Polygenic score for sleep duration. Association with cognition. Sleep Med. 2020;74:262–266. doi:10.1016/j.sleep.2020.07.001

41. Chaput JP, Dutil C, Featherstone R, et al. Sleep duration and health in adults: an overview of systematic reviews. Appl Physiol Nutr Metab. 2020;45(10 (Suppl. 2)):S218–S231. doi:10.1139/apnm-2020-0034

42. Chaput JP, Dutil C, Featherstone R, et al. Sleep timing, sleep consistency, and health in adults: a systematic review. Appl Physiol Nutr Metab. 2020;45(10 (Suppl. 2)):S232–S247. doi:10.1139/apnm-2020-0032

43. Dattani S, Rodés-Guirao L, Ritchie H, Roser M. Mental Health.; 2023.

44. Zelviene P, Kazlauskas E. Adjustment disorder: current perspectives. Neuropsychiatr Dis Treat. 2018;Volume 14:375–381. doi:10.2147/NDT.S121072

45. Freeman D, Sheaves B, Waite F, Harvey AG, Harrison PJ. Sleep disturbance and psychiatric disorders. Lancet Psychiatry. 2020;7(7):628–637. doi:10.1016/S2215-0366(20)30136-X

46. Daghlas I, Lane JM, Saxena R, Vetter C. Genetically Proxied Diurnal Preference, Sleep Timing, and Risk of Major Depressive Disorder. JAMA Psychiatry. 2021;78(8):903. doi:10.1001/jamapsychiatry.2021.0959

47. O’Loughlin J, Casanova F, Jones SE, et al. Using Mendelian Randomisation methods to understand whether diurnal preference is causally related to mental health. Mol Psychiatry. 2021;26(11):6305–6316. doi:10.1038/s41380-021-01157-3

48. Zhou F, Li S, Xu H. Insomnia, sleep duration, and risk of anxiety: A two-sample Mendelian randomization study. J Psychiatr Res. 2022;155:219–225. doi:10.1016/j.jpsychires.2022.08.012

49. Baranova A, Cao H, Zhang F. Shared genetic liability and causal effects between major depressive disorder and insomnia. Hum Mol Genet. 2022;31(8):1336–1345. doi:10.1093/hmg/ddab328

50. Cai L, Bao Y, Fu X, et al. Causal links between major depressive disorder and insomnia: A Mendelian randomisation study. Gene. 2021;768:145271. doi:10.1016/j.gene.2020.145271

51. O’Connell KS, Frei O, Bahrami S, et al. Characterizing the Genetic Overlap Between Psychiatric Disorders and Sleep-Related Phenotypes. Biol Psychiatry. 2021;90(9):621–631. doi:10.1016/j.biopsych.2021.07.007

52. Scott AJ, Webb TL, Martyn-St James M, Rowse G, Weich S. Improving sleep quality leads to better mental health: A meta-analysis of randomised controlled trials. Sleep Med Rev. 2021;60:101556. doi:10.1016/j.smrv.2021.101556

53. Gao X, Meng LX, Ma KL, et al. The bidirectional causal relationships of insomnia with five major psychiatric disorders: A Mendelian randomization study. Eur Psychiatry. 2019;60:79–85. doi:10.1016/j.eurpsy.2019.05.004

54. Hu Y, Shmygelska A, Tran D, Eriksson N, Tung JY, Hinds DA. GWAS of 89,283 individuals identifies genetic variants associated with self-reporting of being a morning person. Nat Commun. 2016;7(1):10448. doi:10.1038/ncomms10448

55. Dashti HS, Ordovás JM. Genetics of Sleep and Insights into Its Relationship with Obesity. Annu Rev Nutr. 2021;41(1):223–252. doi:10.1146/annurev-nutr-082018-124258

56. Ding C, Lim LL, Xu L, Kong APS. Sleep and Obesity. J Obes Metab Syndr. 2018;27(1):4–24. doi:10.7570/jomes.2018.27.1.4

57. Itani O, Jike M, Watanabe N, Kaneita Y. Short sleep duration and health outcomes: a systematic review, meta-analysis, and meta-regression. Sleep Med. 2017;32:246–256. doi:10.1016/j.sleep.2016.08.006

58. Jike M, Itani O, Watanabe N, Buysse DJ, Kaneita Y. Long sleep duration and health outcomes: A systematic review, meta-analysis and meta-regression. Sleep Med Rev. 2018;39:25–36. doi:10.1016/j.smrv.2017.06.011

59. Wu Y, Zhai L, Zhang D. Sleep duration and obesity among adults: a meta-analysis of prospective studies. Sleep Med. 2014;15(12):1456–1462. doi:10.1016/j.sleep.2014.07.018

60. Garfield V. The Association Between Body Mass Index (BMI) and Sleep Duration: Where Are We after nearly Two Decades of Epidemiological Research? Int J Environ Res Public Health. 2019;16(22):4327. doi:10.3390/ijerph16224327

61. Vats MG, Mahboub BH, Al Hariri H, Al Zaabi A, Vats D. Obesity and Sleep-Related Breathing Disorders in Middle East and UAE. Can Respir J. 2016;2016:1–5. doi:10.1155/2016/9673054

62. Hayes BL, Vabistsevits M, Martin RM, Lawlor DA, Richmond RC, Robinson T. Establishing causal relationships between sleep and adiposity traits using Mendelian randomization. Obesity. 2023;31(3):861–870. doi:10.1002/oby.23668

63. Liu X, Li C, Sun X, et al. Genetically Predicted Insomnia in Relation to 14 Cardiovascular Conditions and 17 Cardiometabolic Risk Factors: A Mendelian Randomization Study. J Am Heart Assoc. 2021;10(15):e020187. doi:10.1161/JAHA.120.020187

64. Noordam R, Bos MM, Wang H, et al. Multi-ancestry sleep-by-SNP interaction analysis in 126,926 individuals reveals lipid loci stratified by sleep duration. Nat Commun. 2019;10(1):5121. doi:10.1038/s41467-019-12958-0

65. Wang H, Noordam R, Cade BE, et al. Multi-ancestry genome-wide gene–sleep interactions identify novel loci for blood pressure. Mol Psychiatry. 2021;26(11):6293–6304. doi:10.1038/s41380-021-01087-0

66. Franks PW, Merino J. Gene-lifestyle interplay in type 2 diabetes. Curr Opin Genet Dev. 2018;50:35–40. doi:10.1016/j.gde.2018.02.001

67. Chung J, Goodman M, Huang T, Bertisch S, Redline S. Multidimensional sleep health in a diverse, aging adult cohort: Concepts, advances, and implications for research and intervention. Sleep Health. 2021;7(6):699–707. doi:10.1016/j.sleh.2021.08.005

